# HIV-1 5’-Leader Mutations in Plasma Viruses Before and After the Development of Reverse Transcriptase Inhibitor-Resistance Mutations

**DOI:** 10.1101/2023.06.04.23290942

**Authors:** Janin Nouhin, Philip L. Tzou, Soo-Yon Rhee, Malaya K. Sahoo, Benjamin A. Pinsky, Miri Krupkin, Joseph D. Puglisi, Elisabetta V. Puglisi, Robert W. Shafer

## Abstract

**Background:** HIV-1 RT initiation depends on interaction between viral 5’-leader RNA, RT, and host tRNA3_Lys_. We therefore sought to identify co-evolutionary changes between the 5’-leader and RT in viruses developing RT-inhibitor resistance mutations.

**Methods:** We sequenced 5’-leader positions 37-356 of paired plasma virus samples from 29 individuals developing the NRTI-resistance mutation M184V, 19 developing an NNRTI-resistance mutation, and 32 untreated controls. 5’-leader variants were defined as positions where ≥20% of NGS reads differed from the HXB2 sequence. Emergent mutations were defined as nucleotides undergoing ≥4-fold change in proportion between baseline and follow-up. Mixtures were defined as positions containing ≥2 nucleotides each present in ≥20% of NGS reads.

**Results:** Among 80 baseline sequences, 87 positions (27.2%) contained a variant; 52 contained a mixture. Position 201 was the only position more likely to develop a mutation in the M184V (9/29 vs. 0/32; p=0.0006) or NNRTI-resistance (4/19 vs. 0/32; p=0.02; Fisher’s Exact Test) groups than the control group. Mixtures at positions 200 and 201 occurred in 45.0% and 28.8%, respectively, of baseline samples. Because of the high proportion of mixtures at these positions, we analyzed 5’-leader mixture frequencies in two additional datasets: five publications reporting 294 dideoxyterminator clonal GenBank sequences from 42 individuals and six NCBI BioProjects reporting NGS datasets from 295 individuals. These analyses demonstrated position 200 and 201 mixtures at proportions similar to those in our samples and at frequencies several times higher than at all other 5’-leader positions.

**Conclusions:** Although we did not convincingly document co-evolutionary changes between RT and 5’-leader sequences, we identified a novel phenomenon, wherein positions 200 and 201, immediately downstream of the HIV-1 primer binding site exhibited an extraordinarily high likelihood of containing a nucleotide mixture. Possible explanations for the high mixture rates are that these positions are particularly error-prone or provide a viral fitness advantage.

## INTRODUCTION

The first 356 nucleotides of the RNA genome, the 5’-leader extends from the start of the transactivating response (TAR) element at HXB2 position 455 in the proviral DNA sequence to HXB2 position 810 [1]. The 5’-leader structure and functions have been studied using phylogenetic methods [2], biochemical experiments with site-directed mutants [3–5], RNA structural probing methods [6,7], NMR [8], and cryo-EM [9,10]. These and many other studies have shown that the 5’-leader has multiple functional units including TAR, which is required for initiating transcription; several RNA elements required for RT initiation including the nearly invariant primer binding site (PBS); and several downstream elements responsible for viral splicing, dimerization, and packaging [1,4,8,9,11,12].

The 5’-leader region adapts at least two conformations including a monomeric form that interacts with the cellular translation machinery to favor the synthesis of HIV-1 proteins and a dimeric form that results in genomic packaging and virus assembly (reviewed in [13,14]). The 5’-leader also initiates reverse transcription through a three-way interaction among the viral RNA genome, the reverse transcriptase (RT) enzyme, and tRNA3_Lys_ (reviewed in [4,15]). During RT initiation, RT binds to the viral RNA-tRNA3_Lys_ duplex forming the reverse transcription initiation complex [9].

RT initiation represents a bottleneck to HIV-1 replication as evidenced by an approximately 100-3000-fold reduced rate of nucleotide incorporation during initiation compared with elongation and by the much slower synthesis of the first 200 HIV-1 nucleotides compared to the remaining nucleotides of proviral DNA [15]. We hypothesized that during RT initiation there may be an interplay between changes in RT and the 5’-leader viral RNA. In particular, we questioned whether the 5’-leader would evolve to adapt to the development of drug-resistance mutations (DRMs) in the RT enzyme. Therefore, we sequenced the 5’-leader region of plasma virus samples obtained from individuals before and after the development of the most common nucleoside RT inhibitor (NRTI) mutation M184V, which confers resistance to the cytidine analogs lamivudine (3TC) and emtricitabine (FTC), or a non-nucleoside RT inhibitor (NNRTI)-resistance mutation.

## METHODS

### Individuals and samples

We identified individuals undergoing two or more genotypic resistance tests at different times for clinical purposes who had cryopreserved remnant plasma samples meeting one of the following criteria: (1) Had a baseline sample lacking the 3TC/FTC-resistance mutation M184V/I and a follow-up sample containing M184V (“M184V group”); (2) Had a baseline sample lacking an NNRTI-resistance mutation and a follow-up sample containing an NNRTI-resistance mutation (“NNRTI group”); and (3) Had two or more samples at different times lacking M184V/I or an NNRTI resistance mutation (“Control group”). Individuals meeting the first criteria were required to not have a history of a sample with M184V/I and individuals meeting the second criteria were required to not have a history of a sample with an NNRTI-resistance mutation. To be eligible for next-generation sequencing (NGS), samples were required to have a plasma HIV-1 RNA level ≥10,000 copies/ml.

The remnant plasma samples used in this study were obtained between the years 2000 and 2017. The Stanford University Institutional Review Board approved this study under protocol 6633, entitled “Human Immunodeficiency Virus Quasispecies During Antiviral Therapy”. The approval included a waiver of consent because the study used anonymized de-identified data and remnant plasma samples. The samples were sequenced and the data were collected for research purposes between July 2019 and January 2021.

### Sequencing protocol

Total nucleic acids were extracted from 200 uL of plasma using the EZ1 Virus Mini Kit V2.0 on the automated EZ1 advanced XL (both from Qiagen), according to manufacturer’s instructions. Nucleic acids were eluted in 60uL of AVE buffer and the 5’-leader region was amplified using the following amplification strategy. First, one-step RT-PCR (SuperScript III) was performed using HIV-1 specific primers with 5’ tags to enable sample indexing during a second PCR. The PCR primers were complementary to HXB2 positions 469-490 (5’-GACCAGATCTGAGCCTGGGAGC) and 863-844 (5’-CCCCCTGGCCTTAACCGAAT). The resulting amplicon encompassed HXB2 positions 491-843 which included the 5’-leader positions 37-356. A second PCR was then performed to multiplex samples with dual indexes using NEBNext Multiplex Oligos for Illumina (New England Biolabs). Library quality and concentration were measured using the Agilent DNA 1000 kit (Agilent Technologies). The PCR products showing non-specific peaks were purified again using E-gel SizeSelect II Agarose Gel, 2% (Invitrogen) according to manufacturer’s instructions. Amplicons were pooled at equimolar concentration and purified using AMPure XP beads (Beckman Coulter). The library was spiked with 12.5% PhiX and loaded onto an Illumina MiSeq v2 Cartridge at 8 pM and was sequenced using 2 x 250 bp runs. Both ends of each DNA fragment were sequenced (i.e., paired-end sequencing) to obtain bi-directional sequence information.

### Sequence analysis pipeline

The Fastp program was applied to each FASTQ file to trim adapters, remove regions with low phred scores, and stich paired reads [16]. Trimmed sequence reads were aligned to the HXB2 5’-leader sequence using Minimap2 and the alignments were saved in Sequence Alignment Map (SAM) text files [17]. Samtools were used to convert the resulting SAM text files into binary BAM files and BAI index files [18]. PySam was used to read each BAM file to construct nucleotide frequency files containing the proportions of each nucleotide and indel at each 5’-leader position. Paired samples for which both baseline and follow-up contained a read depth of ≥200 at all 5’-leader positions between 37 and 356 were retained for analysis.

We then created consensus sequences using IUPAC codes, whenever one or more nucleotide was present in ≥20% of sequence reads at the same position. These sequences were submitted to GenBank (accession numbers: OQ814268-OQ814427) and used to construct a neighbor-joining phylogenetic tree. The phylogenetic tree employed a weighted distance matrix, which utilized the Jaccard distance to account for the overlap among nucleotides when more than one nucleotide was present at the same position. The 160 unedited fastq files were submitted to the NCBI Sequence Read Archive (BioProject PRJNA954829).

For each baseline and follow-up sequence, we reported all nucleotides, insertions, and deletions present in ≥20% of sequence reads. Mutational changes between baseline and follow-up were defined as nucleotides or indels for which the proportion of reads containing them changed by ≥4-fold and that were present at follow-up in ≥20% of reads. This requirement therefore occasionally necessitated noting which nucleotides and indels in the baseline sequence were present between 5% and 20% of reads.

### Analysis of previously published HIV-1 group M 5’-leader sequences

#### One-per-person sequence set

We searched the June 2021 version of the Los Alamos National Laboratory HIV Sequence Database (LANL) for non-problematic HIV-1 5’-leader group M sequences with a minimal length of 250 nucleotides. Sequences were grouped into submission sets according to the GenBank “Title” and “Author” fields. Submission sets that were not linked to a PubMed reference or that contained sequences from fewer than five persons were excluded from analysis. When multiple sequences were available for the same person, we randomly selected one sequence for analysis. The search yielded 85 studies containing 1,417 one-per-person 5’-leader sequences.

Each sequence was first aligned to the HXB2 reference 5’-leader sequence using the Smith-Waterman algorithm included in the European Molecular Biology Open Software Suite (EMBOSS) package [19]. Next a multiple sequence alignment of the 5’-leader regions was performed using MAFFT with a set of 157 pre-aligned subtype reference sequences from LANL as a seed alignment using the option for adding unaligned sequences [20]. The alignment was then manually adjusted to increase the consistency of indel placement. For each 5’-leader position we determined the frequency of each nucleotide and each indel.

#### Clonal sequences set

Among the 85 publications reporting HIV-1 group M 5’-leader sequences from ≥5 persons, we identified five publications that reported an average of at least three clones per person with active virus replications [21–25]. Four publications reporting at least three clones per person were excluded because the clones were from proviral DNA samples from persons with virological suppression and they contained large numbers of deletions.

#### NGS set

We reviewed the NCBI Sequence Read Archive (SRA) to identify published studies in which NGS encompassed the 5’-leader in at least five persons. We analyzed the sequences from each of these studies using the NGS pipeline described above (“Sequence analysis section”). Overall, there were 246 SRA BioProjects including 63 containing 5’-leader sequences as determined by our sequence analysis pipeline. Six publications contained sequences from 295 persons (between 8 and 105 persons per publication) [7,26–30].

## RESULTS

### Individuals and samples

Samples before and after ART were available for 29 individuals who developed M184V and for 19 individuals who developed an NNRTI-resistance mutations including K103N (n=13), Y181C (n=5), and G190A (n=1). Samples from two time points were available from 32 control individuals who were ART-naïve and who did not develop any RTI-resistance mutations. Table 1 summarizes the demographics, ART histories, and baseline laboratory values for each group of individuals. Among the 29 individuals in the M184V group, 19 were NRTI-naïve at the time of the first sample. Nine had previously received 3TC or FTC but never developed M184V; one had received NRTIs other than 3TC or FTC. Among the 19 individuals in the NNRTI-resistance group, 16 were NNRTI-naïve and three had previously received an NNRTI but never developed NNRTI resistance.

**Table 1.** Description of the 80 Individuals Undergoing Baseline and Follow-Up HIV-1 5’-Leader Sequencing.

|  |  | Control Group<br>(n=33) | M184V Group<br>(n=29) | NNRTI-Resistance<br>Group<br>(n=19) | P value<sup>1</sup> |  |  |
| --- | --- | --- | --- | --- | --- | --- | --- |
|  |  |  |  |  | Control<br>vs.<br>M184V | Control<br>vs.<br>NNRTI | M184V<br>vs.<br>NNRTI |
| **Sex** |  | 30 male / 2 female | 24 male / 5 female | 16 male / 3 female | NS | NS | NS |
| **Age (IQR)** |  | 39 (26.8-51) | 42 (33-47) | 41 (33-48) | NS | NS | NS |
| **Year (IQR)** |  | 2011 (2006-2014) | 2001 (2001-2004) | 2004 (2001-2010) | <0.001 | 0.003 | NS |
| **Baseline ART history<sup>2</sup>** |  | Naïve: 32 | NRTI-naïve: 19<br>NRTI-experienced: 10 | NNRTI-naïve: 16<br>NNRTI-experienced: 3 | - | - | - |
| **CD4 (IQR)** | *Baseline* | 402.5 (296-503.5) | 83 (35-252) | 160 (78-259) | <0.001 | <0.001 | NS |
|  | *Follow-up* | 315 (221-472.8) | 185 (81-312) | 192 (98-255) | 0.007 | 0.01 | NS |
| **VL (IQR)** | *Baseline* | 4.6 (4.1-4.9) | 5.2 (4.6-5.6) | 4.6 (4.4-5.6) | 0.002 | NS | NS |
|  | *Follow-up* | 4.8 (4.3-5) | 4.4 (4.1-4.8) | 4.8 (4.5-5) | NS | NS | NS |
| **Months (IQR)<sup>3</sup>** |  | 12.5 (2-21.5) | 20 (7-36) | 39 (16.5-54.5) | 0.04 | 0.01 | 0.04 |
| **Pol genetic distance (IQR)<sup>4</sup>** |  | 0.75% (0.44%-1.23%) | 2.17% (1.6%-2.84%) | 1.92% (1.47%-2.98%) | <0.001 | <0.001 | NS |

**Footnotes:** <sup>1</sup>Wilcoxon Rank Sum Tests were used to compare all median values. Fisher's exact tests were used to compare proportions (e.g., proportion that were male). <sup>2</sup>In the M184V Group, 9 of the 10 individuals who were NRTI-experienced individuals at baseline had received a 3TC- or FTC-containing regimen but none had developed M184V (or M184I) prior to initial sampling. In the NNRTI-resistance group, 6 of the 16 individuals who were NNRTI-naïve at baseline were completely ART-naïve; none of the 3 NNRTI-experienced individuals had previous NNRTI-resistance mutations. <sup>3</sup>Months between the baseline and follow-up samples. <sup>4</sup>The percentage of nucleotides that differed between baseline and follow-up in the HIV-1 *pol* sequence performed for clinical purposes at the time the samples were initially obtained. The *pol* sequence usually encompassed the complete protease and the first 300 RT codons.

At the time the initial plasma samples were obtained, the individuals in the control group had a higher median CD4 count (402.5 cells/mm^3^) compared to those in the M184V (83 cells/mm^3^) and NNRTI-resistance (160 cells/mm^3^) groups. At the time of follow-up sampling, the median CD4 count in the control group decreased while the median CD4 count in the M184V and NNRTI groups increased (Table 1). The median time between the two samples differed between the three groups: 12.5 months for the control group, 20 months for the M184V group, and 39 months for the NNRTI-resistance group. The median uncorrected genetic distance in the protease and RT genes was 0.75% for the individuals in the control group compared with 2.17% and 1.92% in the M184V and NNRTI-resistance groups, respectively.

### Baseline sequences

The median number of reads per sample was 2883 (IQR: 1264 – 4981). Within each sample, the coverage was highly uniform across positions 37-356. Across all samples, 98.4% of paired reads encompassed ≥300 nucleotides. All of the sequences belonged to subtype B based on phylogenetic analysis of the pol gene. Among the 80 baseline sequences, 87 (27.2%) positions had a nucleotide difference from HXB2 present in ≥20% of sequence reads (Figure 1). At positions 47, 200, 201, 213, 227, 265, and 305 more than one-half of the nucleotides differed from HXB2 but were the same as the subtype B consensus sequence.

**Figure 1.**
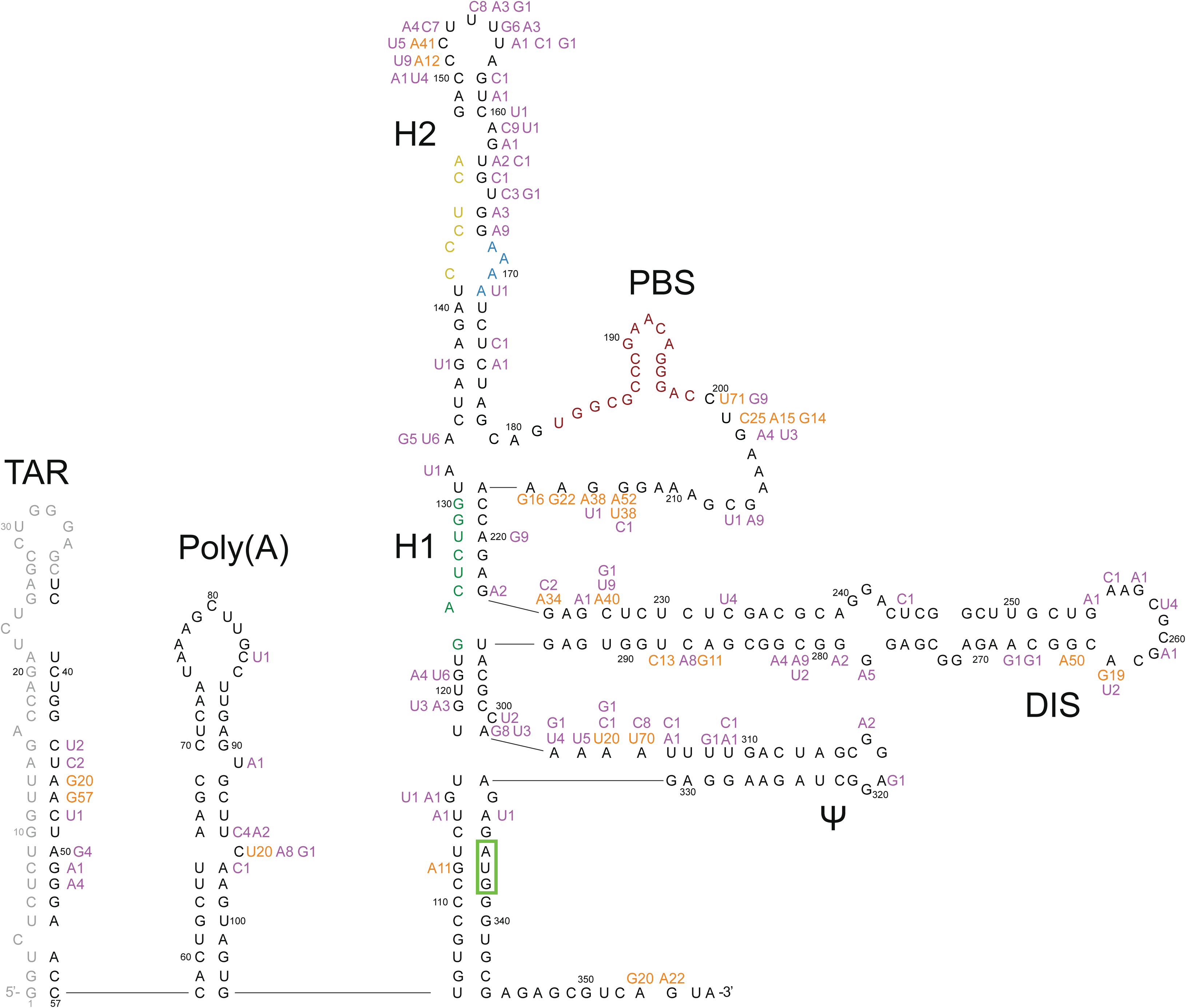
HIV-1 5’-leader nucleotide differences from HXB2 observed in the 80 baseline sequences superimposed on a diagram of its RNA secondary structure. Nucleotides observed in ≥20% of reads in more than 10% of sequences are shown in orange; those present in fewer than 10% of sequences are shown in purple. Insertions and deletions are not shown. Positions 1 to 36 were not sequenced. TAR: trans-activation response element (positions 1-57); PolyA: polyadenylation signal (positions 58-104); H1: helix 1 tertiary structure; H2: helix 2 tertiary structure; PBS: primer binding site (positions 182-199); DIS: dimer initiation site (positions 242-278); psi: packaging signal (positions 312-315). The Matrix initiation codon is surrounded by a green box; it is base-paired to the U5 triplet. The primer activation signal nucleotides are colored green, the C-rich region nucleotides are colored yellow, and the PBS nucleotides are colored maroon.

The poly A motif (AATAAA; positions 73-78), primer activating signal (PAS; positions 125-130) and primer binding site (PBS; positions 182-199) were completely conserved in the 80 baseline sequences. The U5 region was represented by CGT (91% of sequences) or CAT (9% of sequences) each of which maintained complementarity to the Matrix initiation codon (positions 336-338). The dimerization signal (DIS; positions 257-262) was represented by GCGCGC in all but five sequences. The RNA packaging signal (PSI; positions 312-325) contained the same nucleotides in all but three sequences.

Figure 2 shows that the distribution of nucleotides and indels was highly similar between the 80 baseline samples and the 1,417 samples downloaded from the LANL database (Supplementary Figure 1). There were no apparent differences in the proportions of nucleotides or indels between the sequences from the 56 individuals who were ART-naïve and the 24 individuals who were ART-experienced at baseline (Supplementary Figure 2). Figure 3 shows that 52 positions had more than one nucleotide detected in ≥20% of sequence reads. At positions 200, 201, 304, 224, and 354, more than 10% of samples had a mixture of two nucleotides.

**Figure 2.**
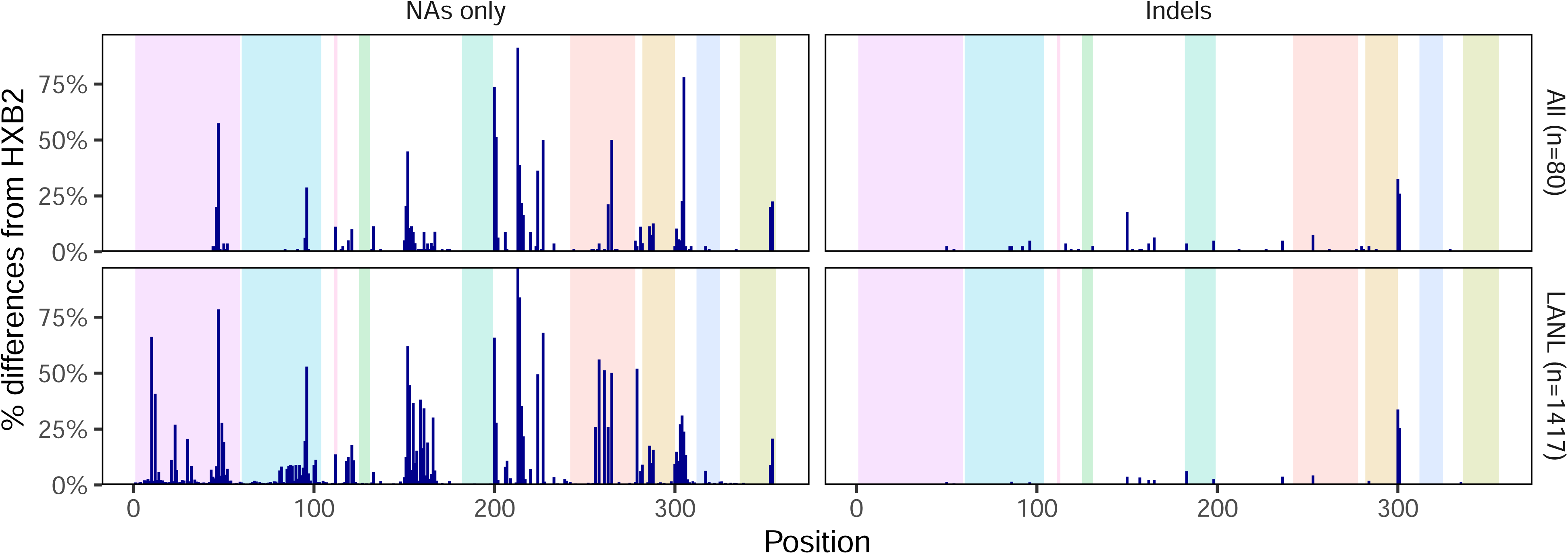
Distribution of HIV-1 5’-leader nucleotide differences from HXB2 and indels in the 80 baseline sequences and in 1,417 one-per-person Los Alamos National Laboratory HIV Sequence Database (LANL) dataset.

**Figure 3.**
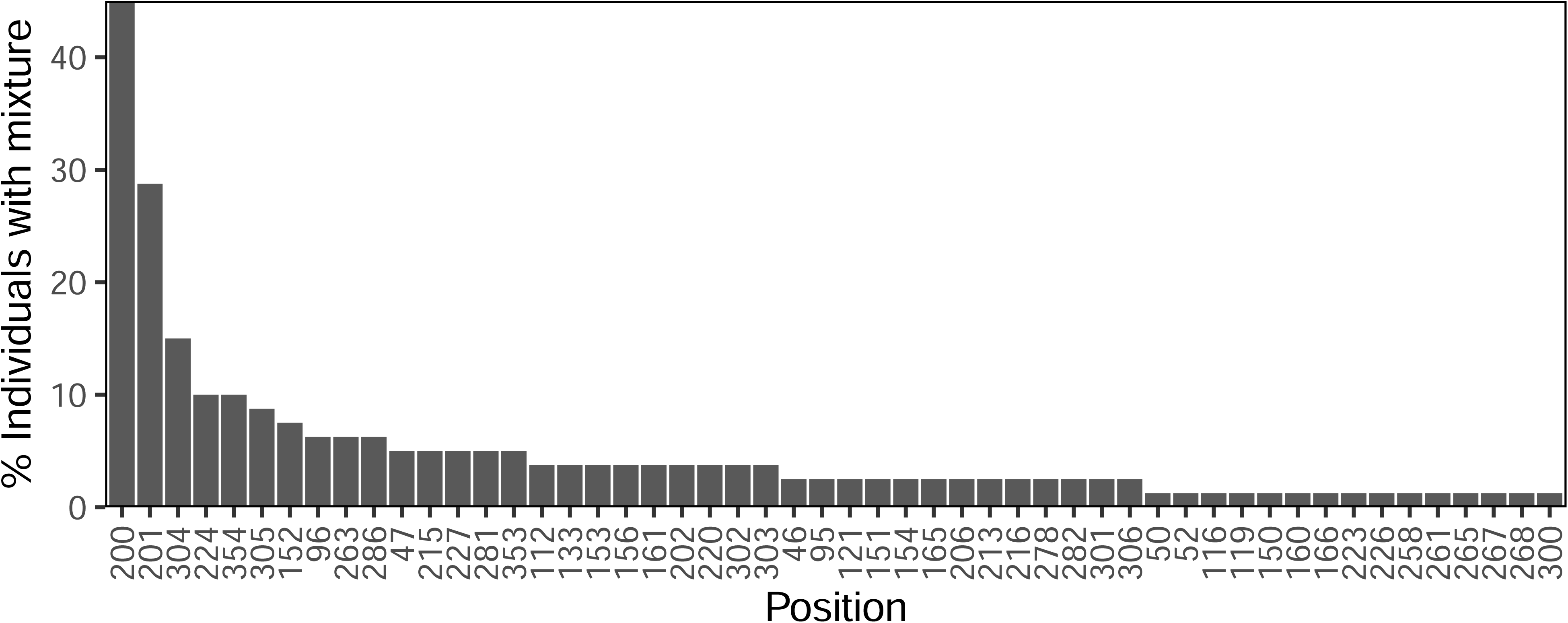
HIV-1 5’-leader positions ranked according to the frequency with which they contained a mixture of two or more nucleotides. Each nucleotide in a mixture was required to present in ≥20% of NGS reads.

Among the 56 individuals who were ART-naïve at the time baseline sequencing was performed, 22 were found by Sanger sequencing of the pol gene to have ambiguous nucleotides (i.e., mixtures) at <0.5% of positions, an established marker suggestive of recent infection [31,32] while 34 were found to have ambiguous nucleotides at ≥0.5% of positions. The probabilities of having mixtures at positions 200 and 201 were significantly correlated with the proportion of ambiguous nucleotides in pol (position 200: spearman rho=0.28, p=0.03; position 201: spearman rho=0.33; p=0.01).

Compared to HXB2, two individuals had large deletions. In one individual, positions 119 to 161 were deleted and in the other individual, positions 281 to 288 were deleted. Both of these deletions were also present in the follow-up sequence from the same individual suggesting that these deletions were not sequencing artifacts. An additional 42 baseline samples had deletions of 1 to 3 nucleotides. The most frequent positions with deletions were between positions 301 to 303 (20 samples) and between positions 150 to 153 (13 samples). Compared to HXB2, 54 samples had one or more insertions, with the most frequent occurring between positions 300 to 305 (26 samples), 254 to 255 (6 samples), 165 to 167 (5 samples), and 198 to 202 (4 samples). Eight samples had insertions containing more than three nucleotides.

### Follow-up sequences

Figure 4 summarizes the number of individuals by the number of positions at which a ≥4-fold change in the proportion of a nucleotide or indel occurred between baseline and follow-up for each of the three sample groups. The median number of changes per individual was higher in the M184V (3; IQR:1-4; p=0.014 Wilcoxon Rank Sum Test) and NNRTI groups (2; IQR: 1-4; p=0.12; Wilcoxon Rank Sum Test) than in the control group (0.5; IQR:0-4). The three control group individuals with large changes included one individual who may have experienced a re-infection as evidenced by a nucleotide distance of 5.4% between their baseline and follow-up HIV-1 pol sequence.

**Figure 4.**
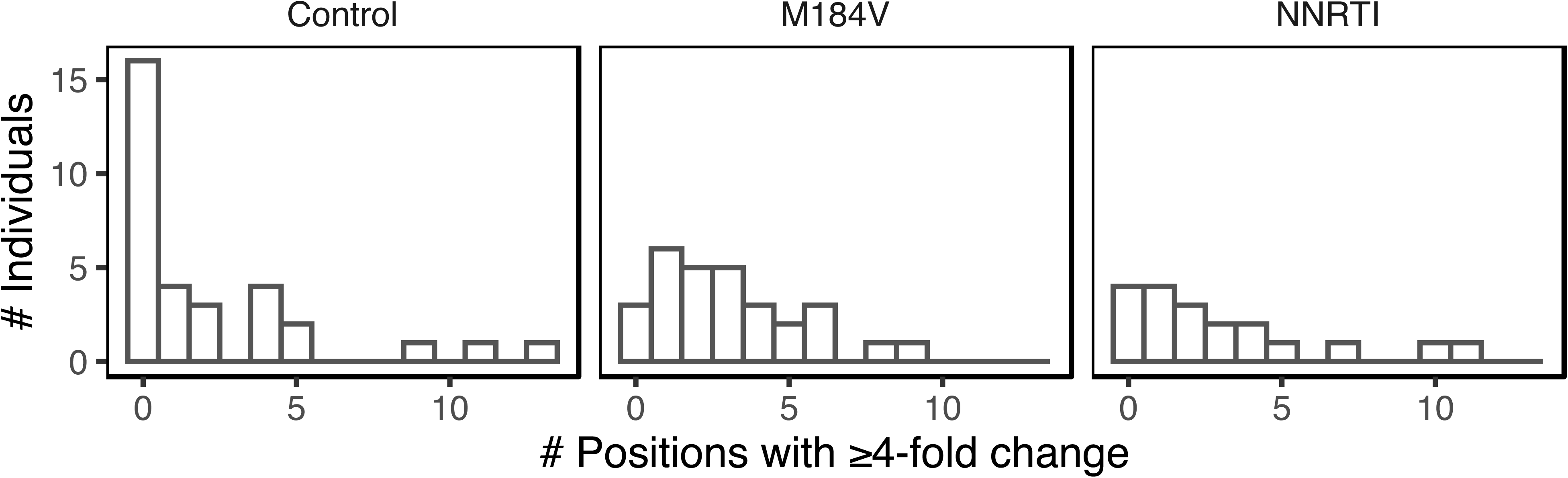
Number of HIV-1 5’-leader positions at which a nucleotide or indel exhibited a ≥4-fold change in its proportion between baseline and follow-up and which was present at a level of ≥20% at follow-up in each of the three patient groups.

Fifty-three of the 63 deletions found in either baseline or follow-up samples were present at both time points. Fifty-three of the 70 insertions present in either baseline or follow-up samples were present at both time points. A neighbor-joining phylogenetic tree using the Jaccard distance to weight the distance between overlapping IUPAC nucleotides is shown in Figure 5.

**Figure 5.**
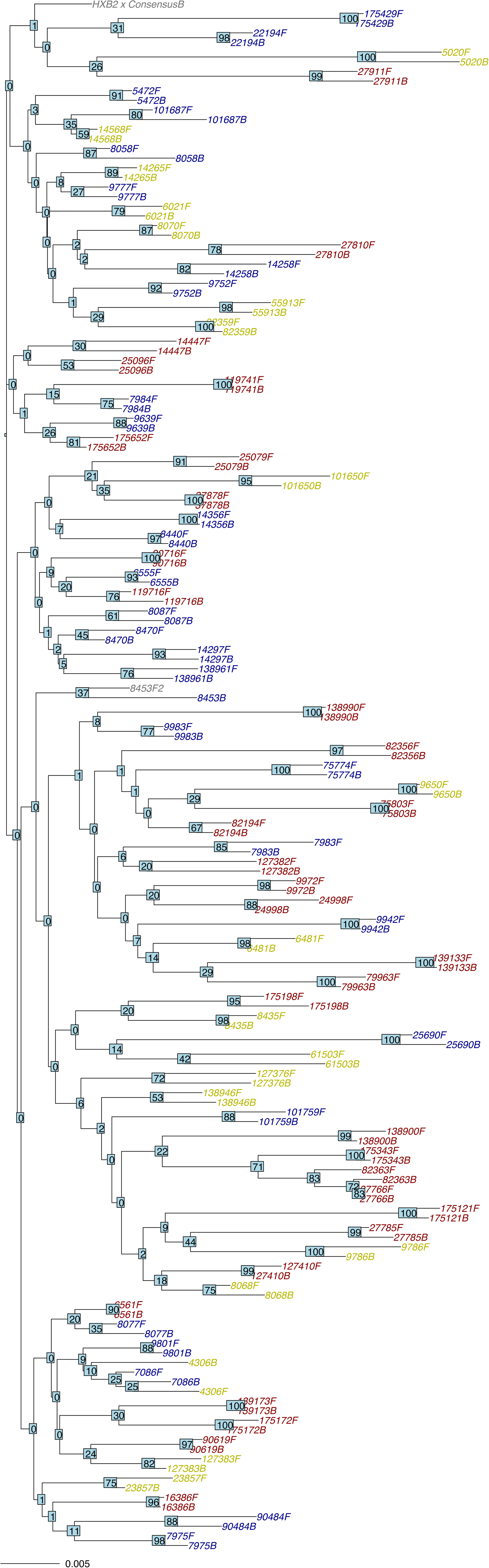
Neighbor-joining tree containing the 80 baseline and follow-up sequences from this study. Sample IDs for the M184V group are colored blue; those for the NNRTI group are colored yellow; and those for the Control group are colored red. Bootstrap values are indicated at each node of the tree.

Figure 6A shows the positions at which at least one nucleotide changed its proportion by ≥4-fold and which occurred in ≥20% of sequence reads. The positions at which nucleotide changes occurred most frequently are shown in Figure 6B. Positions 200 and 201, which were the positions most likely to have mixtures at baseline, were also the positions most likely to exhibit a ≥4-fold change in the proportion of a nucleotide. Position 201 was significantly more likely to exhibit a ≥4-fold change in its proportions in the M184V (9/29 vs. 0/32; p=0.0006; Fisher’s Exact Test) and the NNRTI-resistance groups (4/19 vs. 0/32; p=0.02; Fisher’s Exact Test) compared with the control group. There were no significant differences between each of the three groups at any other position including position 200.

**Figure 6.**
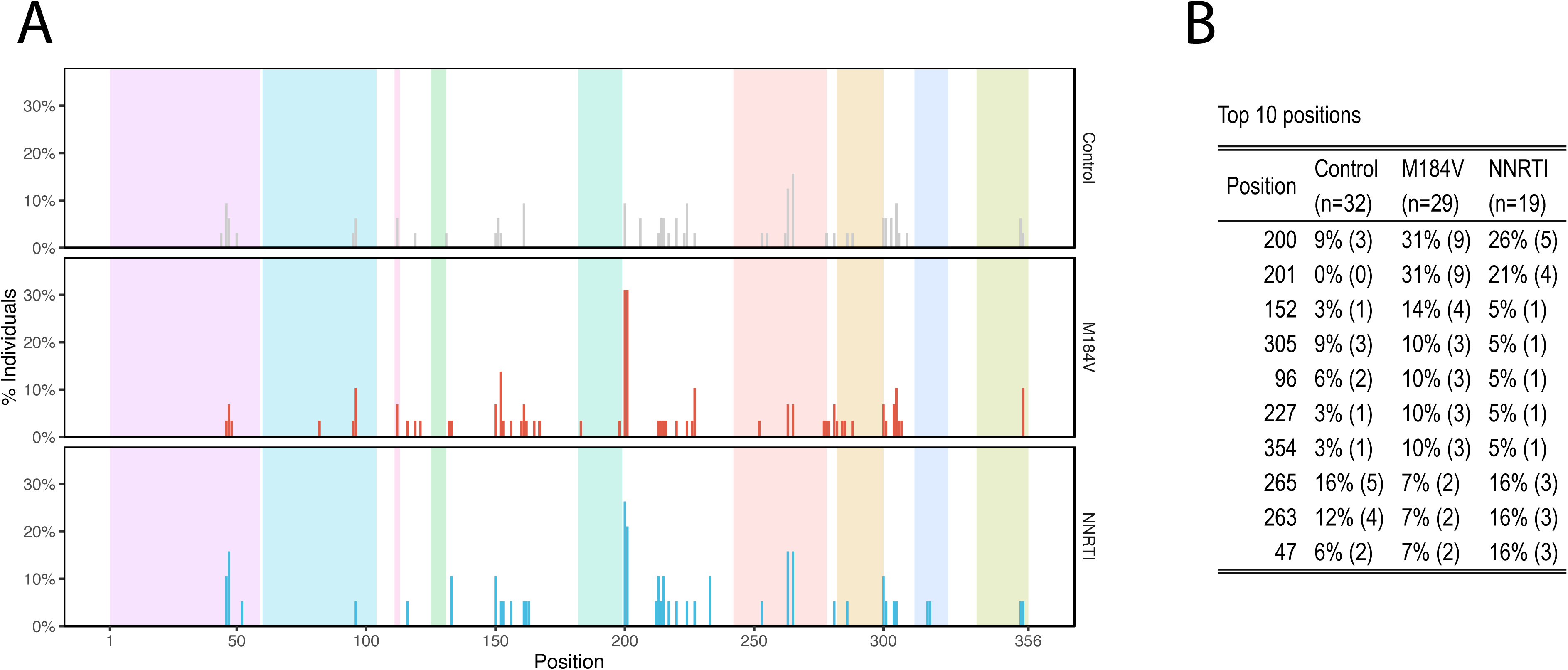
Distribution of positions at which a nucleotide or indel changed its prevalence by ≥4-fold according to patient group (A) and a table indicating the ten positions that changed most often (B).

Although position 201 frequently displayed a change between baseline and follow-up in the M184V and NNRTI groups, there was no consistent pattern of nucleotide change. For example, among the nine individuals in the M184V group that experienced a 4-fold change in the proportion of a nucleotide, the baseline nucleotides were C in five individuals, T in three individuals, and G in one individual. Among the five individuals with C at baseline, four changed to T and one to G. Among the three individuals with T at baseline, one changed to C, another to G, and the third to G and A. The one individual with G changed to T.

### Distribution of mixtures in previously published clonal sequences and in NGS

The five publications describing a minimum of three clones per individual from at least five individuals reported a total of 294 sequences from 42 individuals. At five positions, ≥10% of individuals had a mixture of ≥2 nucleotides in different clones including positions 200 (n=16 individuals; 38.1%), 201 (n=11 individuals; 26.2%), 305 (n=7 individuals; 16.7%), 152 (n=5 individuals; 11.9%), and 281 (n=5 individuals; 11.9%).

The six publications in the NCBI SRA containing 5’-leader sequences from 295 individuals reported that the most common positions containing a mixture with at least two nucleotides (i.e., each present in ≥20% of NGS reads) were also positions 200 (n=103 individuals; 34.9%) and 201 (n=62 individuals; 21.1%). Table 2 illustrates the 15 positions that most frequently contained mixtures in this study and the frequency with which these positions contained mixtures in the GenBank and NCBI SRA datasets. In contrast to the 5’-leader, among 384 RT sequences from 379 individuals in the same six NCBI SRA dataset publications, no position had a mixture in ≥6% of positions.

**Table 2.** Proportion of HIV-1 5’-Leader Positions Containing a Mixture of Nucleotides in Sampled Viruses in this Study, in Previously Published Studies in GenBank, and in NGS Studies in the NCBI Sequence Read Archive (SRA)

| **Position** | **Baseline Samples from this Study**<br>**(n=80 individuals)<sup>1</sup>** | **Published Studies of Individuals with Multiple Clones (GenBank)**<br>**(n=42 individuals)<sup>2</sup>** | **Published Studies of Individuals Undergoing NGS (NCBI SRA)**<br>**(n=295 individuals)<sup>3</sup>** |
| --- | --- | --- | --- |
| 200 | 45.0% | 38.1% | 34.9% |
| 201 | 28.8% | 26.2% | 21.1% |
| 304 | 15.0% | 4.8% | 4.1% |
| 224 | 10.0% | 9.5% | 6.1% |
| 354 | 10.0% | 2.4% | 1.4% |
| 305 | 8.8% | 16.7% | 6.0% |
| 152 | 7.5% | 11.9% | 6.8% |
| 286 | 6.3% | 9.5% | 5.8% |
| 96 | 6.3% | 6.9% | 8.2% |
| 263 | 6.3% | 2.4% | 2.7% |
| 281 | 5.0% | 11.9% | 1.5% |
| 47 | 5.0% | 4.8% | 8.2% |
| 215 | 5.0% | 7.1% | 2.5% |
| 227 | 5.0% | 4.8% | 2.7% |
| 353 | 5.0% | 2.4% | 1.0% |

Footnote: <sup>1</sup>A position was considered to have a mixture if at least one non-consensus nucleotide was present in $\geq 20\%$ of reads. <sup>2</sup>Data were obtained from five publications reporting $\geq 3$ clones per individual from $\geq 5$ individuals. A total of 294 sequences were performed (i.e., a mean of 7 clones per individual). A position was considered to have a mixture if at least one clone had a non-consensus nucleotide. Thirty-one individuals (74%) had subtype B viruses and 11 patients had subtype C viruses. <sup>3</sup>A position was considered to have a mixture if at least one non-consensus nucleotide was present in $\geq 20\%$ of reads. Sixty-four percent of sequences belonged to subtype C; 21% to subtype B; and 15% to other subtypes or circulating recombinant forms.

## DISCUSSION

To our knowledge, this is one of the largest studies of 5’-leader sequences in persons living with HIV-1 and the first study of paired HIV-1 5’-leader sequences from individuals who did or did not develop RT-associated DRMs. The fact that all of the samples were from plasma suggests that the observed variants were most likely replication competent, which is much less often the case for proviral HIV-1 DNA sequences obtained from peripheral blood mononuclear cells (PBMCs) [33]. The availability of sequences from two time points also provides confirmation that the few observed uncommon variants, such as large deletions, were consistent with virus replication.

Contrary to our initial hypothesis, we did not observe co-evolutionary changes between RT and 5’-leader sequences. The only 5’-leader position (position 201) that demonstrated a higher likelihood of evolution in samples from individuals who developed RT-associated DRMs is one of the most variable 5’-leader positions. However, our study uncovered a previously unreported phenomenon, wherein specific 5’-leader positions, particularly positions 200 and 201, exhibited an extraordinarily high likelihood of containing two or more nucleotides each in more than 20% of NGS reads. We corroborated this finding by analyzing two previously published datasets: five published studies of clonal dideoxy-terminator Sanger sequences and six NCBI SRA NGS datasets encompassing the HIV-1 5’-leader.

Although HIV-1 is a quasispecies characterized by the presence of multiple circulating variants, the markedly high prevalence of nucleotide mixtures at two particular positions is striking. The prevalence of mixtures in our 80 baseline sequences was 45% for position 200 and 29% for position 201. In our analysis of previously published clonal sequences, these prevalences were 38% and 26%, respectively, and in our analysis of previously published NGS data, these prevalences were 35% and 21%, respectively. Moreover, in these analyses, positions 200 and 201 contained mixtures at a frequency several times higher than that observed at any other 5’-leader position or, in the NCBI SRA NGS dataset, at any RT position. The fact that the previously published clonal and NGS datasets contained large numbers of non-subtype B isolates suggests that the propensity of positions 200 and 201 to contain nucleotide mixtures is not limited to one subtype.

Positions 200 and 201 are immediately downstream of the PBS, situated opposite to the consecutive A nucleotides at tRNA positions 58 and 57, respectively [9]. The significance of complementarity between the 5’-leader and tRNA at these two positions, however, has not been studied. There are several plausible non-exclusive explanations for the high proportion of mixtures at these two positions. First, these positions may be particularly error prone as they represent the first two nucleotides added following the second strand transfer step during HIV-1 reverse transcription [15]. Second, the presence of multiple circulating variants at these positions may provide an as yet unknown fitness advantage within an individual patient. Third, HIV-1 replication has been shown to be occasionally primed by tRNA5_Lys_, as well as tRNA3_Lys_ [34]; but it is not known whether or how this might influence the development of mutations just following the PBS. Finally, if the tRNA molecules co-packaged with genomic RNA re-annealed to the PBS following our nucleic acid extraction procedure but before reverse transcription, this might also influence mutations just following the PBS.

Our study has several limitations. First, our sequences did not encompass the first 36 nucleotides of TAR. The plasma virus samples that we sequenced begin at the start of TAR and could not be amplified without designing primers that bind to this region making it impossible to sequence the nucleotides bound to or upstream of our 5’-prime PCR primer. Several studies have reported that the 5’-leader structure is exquisitely sensitive to mutations in TAR [35–37]. Indeed, a single nucleotide difference in the transcription start of TAR has been found to influence the overall structure of the 5’-leader [38].

Second, our study is also limited because we studied convenience samples. As a result, we were unable to match the three groups of patients for several important characteristics such as year of infection, time between samples, and CD4 count. However, because we found few differences in the evolution of 5’-leader sequence between the three groups of patients and because our main finding, the extraordinary high proportion of mixtures at positions 200 and 201, was apparent in the baseline sequences, this limitation is unlikely to have influenced our study’s conclusions.

Third, M184V has been shown to increase the fidelity of the HIV-1 RT enzyme [39,40]. Thus, we cannot be certain that the number of mutations in the M184V group was influenced by the development of this mutation. Nonetheless, M184V does not appear to significantly limit the evolutionary potential of viruses containing this mutation in vitro and likely in vivo [41–43]. Finally, we performed NGS without using unique molecular identifiers (UMIs) [44]. The use of UMIs would have allowed us to precisely quantify each variant within the HIV-1 quasispecies and to thus reliably identify linkages between positions in the same variant [44].

In conclusion, this is the first study in which paired circulating plasma HIV-1 5’-leader sequences were obtained from a large number of individuals with well characterized ARV treatment histories at two time points. The study uncovered a previously unreported phenomenon in which several 5’-leader positions, particularly positions 200 and 201, which are immediately downstream of the PBS, often contain high proportions of different nucleotides in the same sample. Data from this study also suggests that nucleotide mixtures at these positions increase with the duration of infection. Future studies employing deep sequencing will provide more insight into this phenomenon by determining whether different nucleotides at positions 200 and 201 are associated with changes at other 5’-leader positions. Additionally, mutational studies will be required to determine the potential biological significance of variations at these positions.

## Data Availability

All data produced are available online at Genbank (accession numbers: OQ814268-OQ814427) and NCBI Sequence Read Archive (BioProject PRJNA954829).

https://www.ncbi.nlm.nih.gov/bioproject/?term=PRJNA954829

## Acknowledgements

JN and RWS have been funded in part by an NIH grant R03 AI147632. EVP, JDP and MK have been funded in part by an NIH grant U54 AI170856.

## Conflicts of interest

The author(s) declare that there are no conflicts of interest.

## FIGURE LEGENDS

**Supplementary Figure 1.** HIV-1 5-leader nucleotide differences from the HXB2 sequence observed in 1,417 sequences in the Los Alamos National Laboratories HIV Sequence Database dataset. Superscripts indicate the percentage of times that a nucleotide was reported. Highlighted regions include (1) TAR: trans-activation response element; (2) PolyA: polyadenylation signal loop with a box surrounding the poly A motif; (3) U5; (4) PAS: primer activation signal; (5) PBS: primer binding site; (6) DIS: dimer initiation signal loop with a box surrounding the palindromic DIS; (7) major 5’ splicing site; (8) PSI packaging signal; (9) Matrix protein with a box surrounding the start codon.

**Supplementary Figure 2.** Distribution of HIV-1 5’-leader nucleotide differences from HXB2 and indels in the 56 baseline sequences from ART-naïve individuals compared with the 24 baseline sequences from ART-experienced individuals.

## Notes

### Competing Interest Statement

The authors have declared no competing interest.

### Author Declarations

The Stanford University Institutional Review Board approved this study under protocol 6633, entitled “Human Immunodeficiency Virus Quasispecies During Antiviral Therapy”.

### Summary of Updates

We performed an additional analysis which we believe strengthens the manuscript. A few more modifications were applied to the manuscript.

