## Supplementary figures and images for "HIV-1 5’-Leader Mutations in Plasma Viruses Before and After the Development of Reverse Transcriptase Inhibitor-Resistance Mutations"

### Supplementary Figure 1

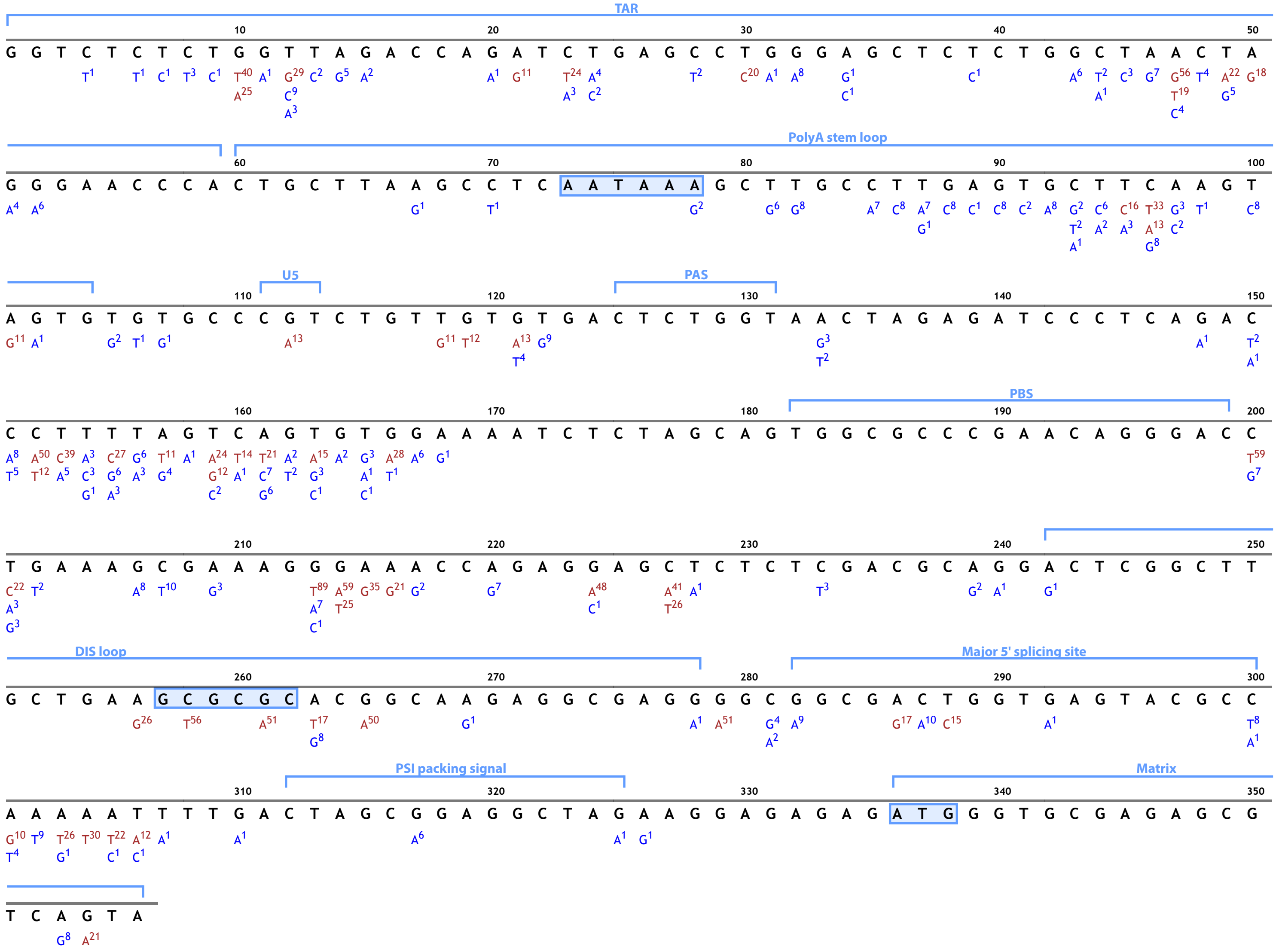

### Supplementary Figure 2

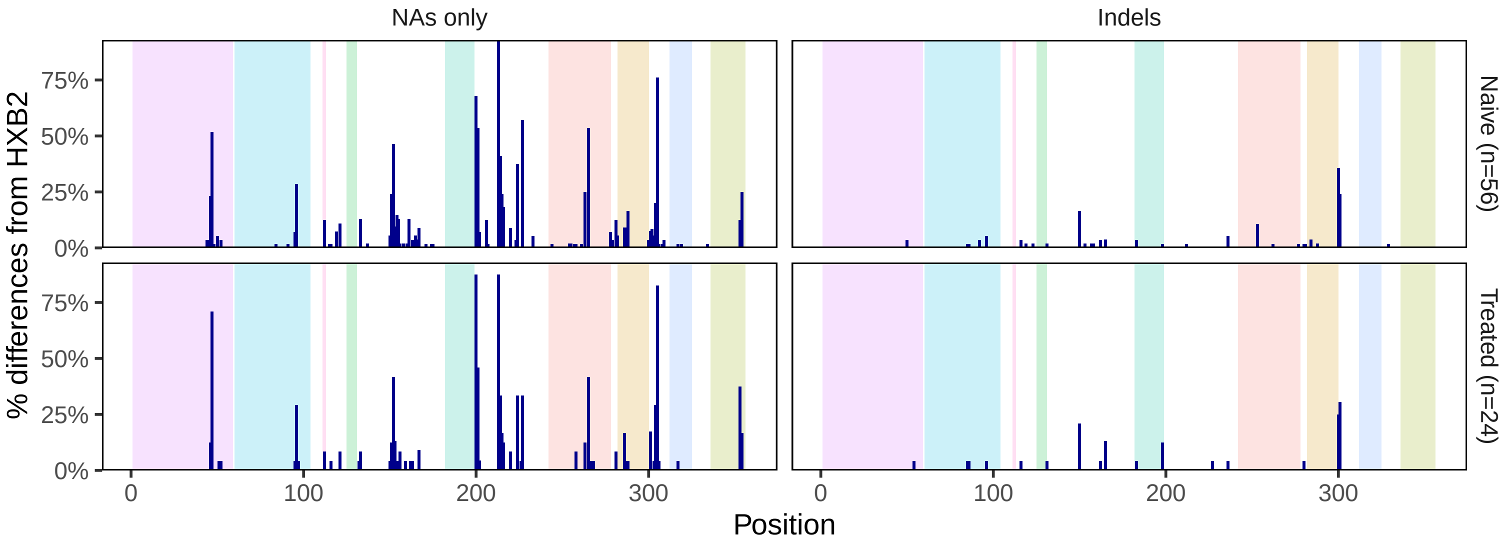
